# Integration of Mendelian Randomization and Genome-wide Association Analysis Reveals Potential Benefits of Cheese Intake on Human Intelligence

**DOI:** 10.1101/2024.10.15.24315561

**Authors:** Peng Chen, Zhikang Cao, Jing Feng, Zhipeng Li, Shaoping Nie

**Affiliations:** State Key Laboratory of Food Science and Resources, Nanchang University, 235 Nanjing East Road, Nanchang 330047, China

## Abstract

The brain relies on a range of nutrients for optimal functioning. However, the causal nature between dietary components and human intelligence are still unclear. This study integrates Mendelian randomization (MR) and genome-wide associations analyses to investigate the relationships. The results show that cheese exerts the most remarkable genetic correlation and causal association with cognition-related traits. We conduct a large-scale meta-analysis of genome-wide associations studies (GWAS) consisting of over 800,000 participants and discover 71 genetic determinants for cheese consumption. Additional MR using the GWAS results recovers the causal effects of cheese on all the cognition-related traits, and also reveals its effects on education, psychological states and gene expressions in multiple brain tissues. Additionally, knockout mouse models and pathway enrichment analysis indicate that the loci mapped genes are involved in cognitive functions and brain characteristics. In conclusion, our comprehensive analysis supports the potential benefits of cheese consumption on human intelligence.

## Introduction

Intelligence is divided into crystallized intelligence and fluid intelligence(*1*). The former includes the ability to use knowledge gained from past experiences to solve problems or understand concepts(*2*). The latter, in contrast, refers to the capacity to solve new and unfamiliar reasoning problems(*3*). Intelligence, particularly fluid intelligence, is significantly influenced by genetic factors. Although 30%-70% of the variance in intelligence quotient may be hereditary(*4*), studies have shown that it can also be affected by environmental factors such as diet, health status, and physical activity(*5*). Diet, in particular, has a significant impact on human intelligence. The effects of diet are mediated through various mechanisms, including nutritional supply, metabolic regulation, neuroprotection, and support of cognitive functions.

A balanced diet is linked to improved cognitive performance on verbal, visuospatial, and memory tasks for diverse age groups, while malnutrition or inadequate nutrition can lead to declines in neurocognitive functions(*6*). A diet that emphasizes whole grains, lean proteins, fruits, and vegetables, often characterized as the Mediterranean diet, has been associated with a lower risk of cognitive decline and a reduced incidence of neurodegenerative diseases in various populations(*7*). Especially, protein is an essential material foundation for brain development, and a deficiency in protein directly affects the structure and function of the brain. Epidemiological evidence has been shown that dairy product consumption may prevent cognitive decline. For example, increased cheese consumption reduces the risk of cognitive impairment by 32%, while milk intake does not alter the risk(*8*). Both animal and human studies have shown that casein peptide, rich in cheese, has positive effects on cognitive functions(*9*).

Indeed, many dietary components have been studied for their potential in enhancing cognitive functions. For example, β-glucan, a form of soluble dietary fiber, has been reported to attenuate cognitive impairment via the gut-brain axis in diet-induced obese mice(*10*). Blueberry extract, rich in anthocyanins, has been shown to exert neuroprotective effects and improve cognitive performance in animal studies(*11*). Increased consumption of nuts and legumes has been linked to better cognitive performance(*12*). The intake of 𝜔-3 fatty acids, commonly found in fish, has also been linked to improvements in brain health. These fatty acids are essential components of neuronal membranes(*13*) and have been shown to play a role in neurogenesis, neuroplasticity, and protection against oxidative stress(*14*). Furthermore, lecithin, abundant in eggs and soybeans, is a source of choline, which serves as a precursor for the synthesis of acetylcholine, a key neurotransmitter(*15*). Phosphatidylserine, found in meat, fish, and beans, is involved in multiple functions of the brain, including activation of membrane signaling pathways, neuroinflammation, neurotransmission, and synaptic refinement(*16*).

Specific dietary components not only support general health but also enhance cognitive functions and provide a buffer against the development of neurodegenerative diseases. As such, diet is an important modifiable risk factor for maintaining cognitive health throughout the lifespan. Although the correlations between many dietary components and cognitive functions have been suggested, the causal nature of these correlations in humans are not clear.

In this work, we employed linkage disequilibrium (LD) score regression and Mendelian Randomization (MR) analysis to explore the relationships between seventeen commonly consumed dietary components and cognition-related traits, including cognitive performance, intelligence, childhood intelligence, fluid intelligence questions attempted within time limit, and fluid intelligence scores (**Fig. 1**). Cheese intake showed the strongest positive causal relationships with all these traits. Subsequently, our meta-analysis of large-scale genome-wide associations studies (GWAS) data including more than 800 thousand subjects identified novel genetic loci strongly associated with cheese consumption. Positional and eQTL mapping of these loci identified genes that play a relevant role in brain functions. Further MR using the GWAS results not only confirmed the causal effects of cheese intake on all the cognition-related traits, but also revealed its positive influence on education, mental states, depression, and anxiety. Moreover, our eQTL-based MR on cheese intake and gene expression in twelve brain tissues provided additional insights in the potential benefits of cheese on cognitive health.

**Fig 1.**
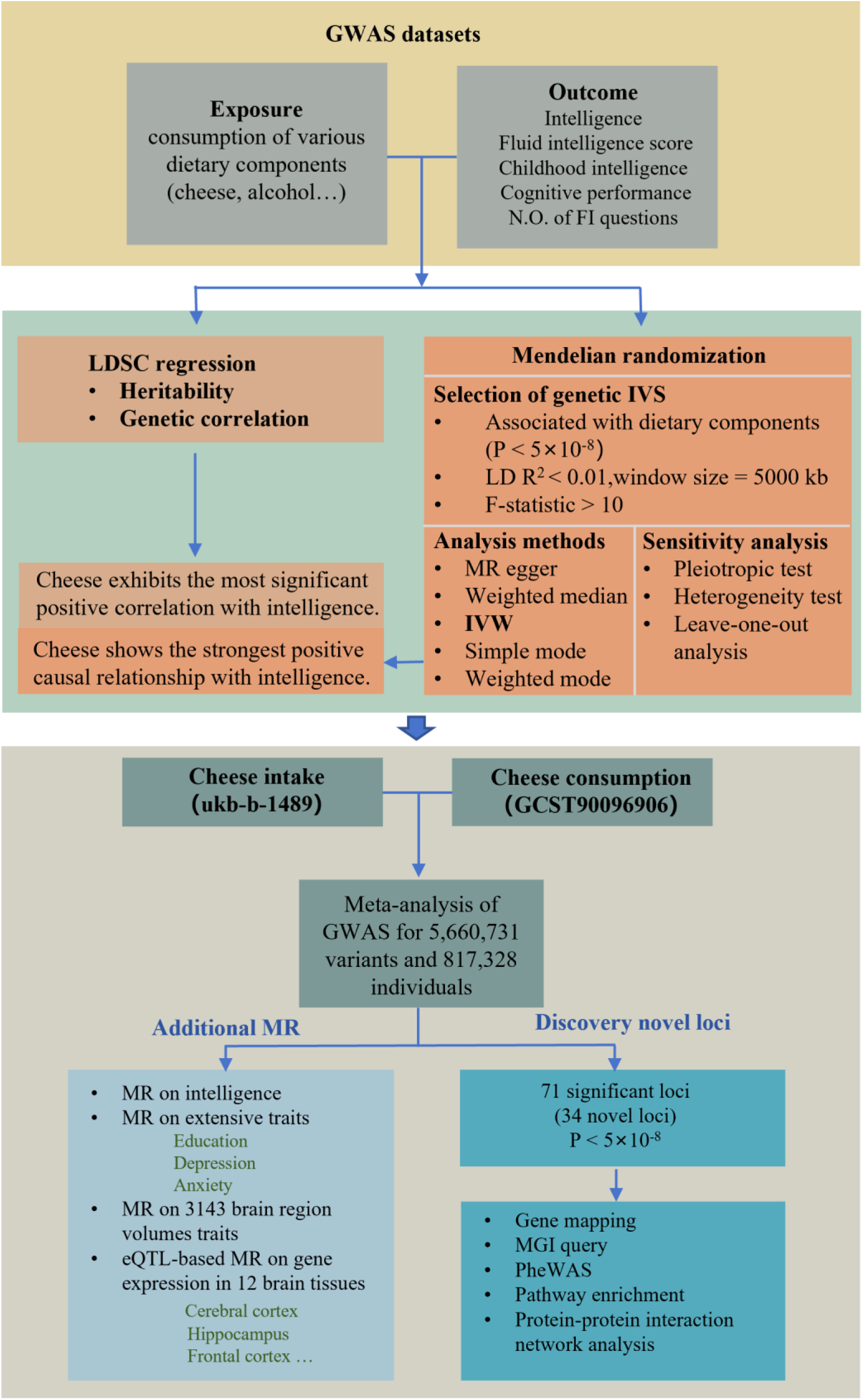
Schematic diagram of the datasets and analyses. FI fluid intelligence, LDSC Linkage Disequilibrium Score Regression, MR Mendelian randomization, eQTL Expression Quantitative Trait Loci, MGI Mouse Genome Informatics, IVW Inverse Variance-Weighted.

## Results

### Genetic correlation between the consumption of dietary components and human intelligence

Firstly, we used LD score regression to determine the heritability of human intelligence. As expected, traits related to intelligence and cognitive functions are highly heritable. Specifically, cognitive performance shows a heritability of 19.90% (FDR = 1.66×10^−181^), intelligence at 18.59% (FDR = 4.78×10^−147^), fluid intelligence score at 22.13% (FDR = 2.13×10^−125^), the number of fluid intelligence questions attempted within the time limit at 11.09% (FDR = 3.29×10^−65^), and childhood intelligence at 27.52% (FDR = 1.19×10^−9^), as illustrated in **Fig. 2A**. We further examined the impacts of genetic factors on dietary preference. Interestingly, the analysis shows that the consumption of all the dietary components explored is significantly influenced by genetic factors, with heritability levels ranging from 2% to 8%. For example, genetic differences account for 6.74% of the variation in cheese consumption (FDR=2.95×10^−134^), while genetics explains 8.02% of the variation in alcohol preference (FDR=5.61×10^−117^) (**Fig. 2B**). Interestingly, the traits related to cognition demonstrated stronger heritability than the dietary intakes, which aligns with our intuition.

**Fig. 2.**
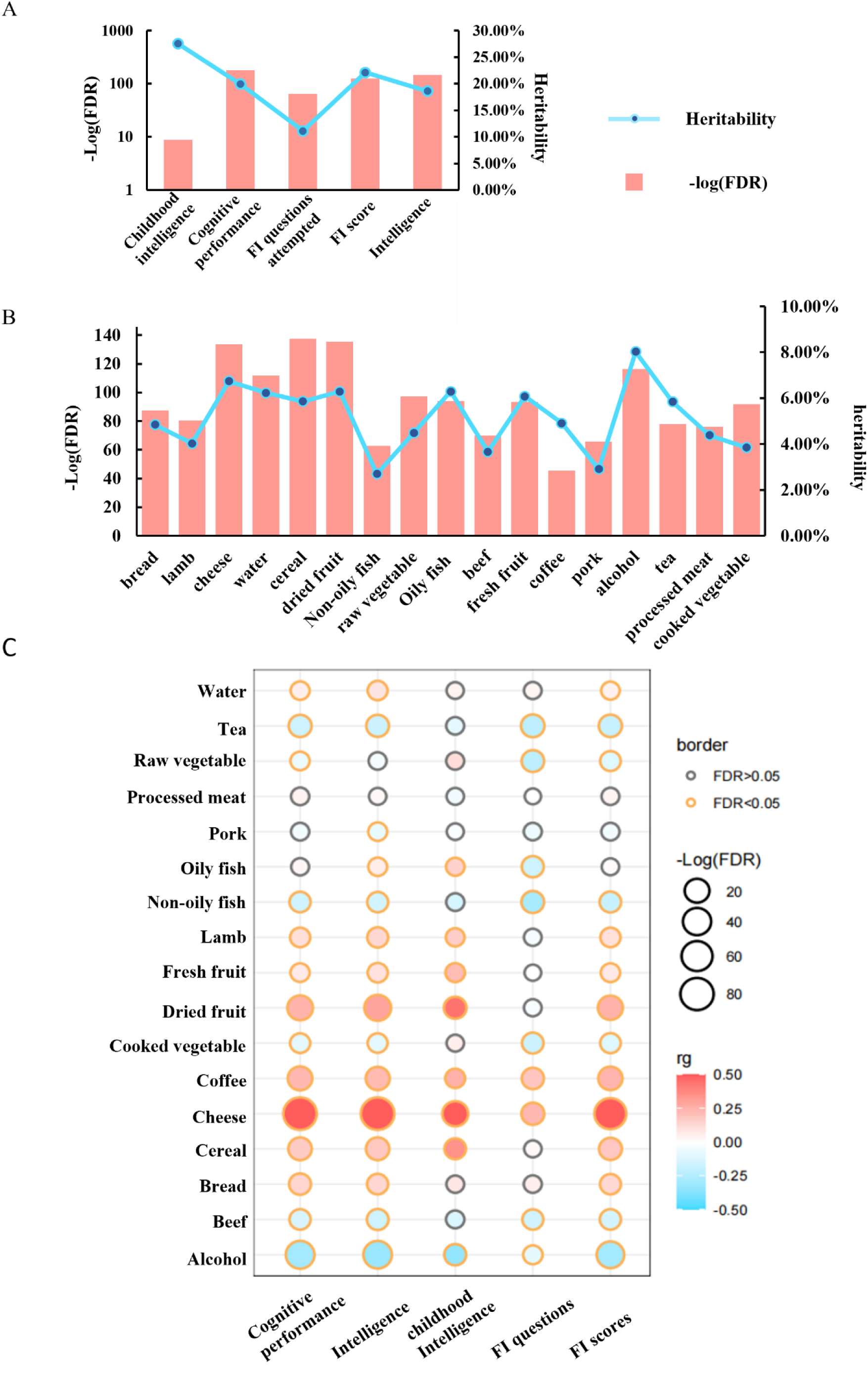
Linkage Disequilibrium score regression analysis of the consumption of various dietary components and phenotypes related cognitive functions. A-B The heritability of cognitive functions and various dietary intakes; C, The genetic correlations between various dietary intakes and cognitive functions. The color of the bubbles corresponds to the genetic associations. The size of the bubbles corresponds to the negative logarithm of the association FDR. All tests were adjusted for multiple comparisons, and associations passing the threshold of FDR<0.05 are indicated by a yellow border.

We proceeded to examine the genetic correlations between these components and cognitive health. The results revealed significant correlations between multiple dietary components and brain function (**Fig. 2C**). Notably, dried fruit exhibits a strong positive correlation with cognitive performance (FDR=1.02×10^−24^), intelligence (FDR=5.78×10^−32^), childhood intelligence (FDR=3.04×10^−12^), and fluid intelligence scores (FDR=1.12×10^−22^). Similarly, coffee is positively correlated with cognitive performance (FDR=1.95×10^−20^), intelligence (FDR=2.88×10^−17^), childhood intelligence (FDR=1.14×10^−4^), the number of fluid intelligence questions attempted (FDR=3.67×10^−10^), and fluid intelligence scores (FDR=3.55×10^−18^). Cheese and alcohol, among the seventeen dietary components analyzed, exhibited the most striking correlations across all traits examined. Cheese positively affects cognitive performance (FDR=1.52×10^−84^), intelligence (FDR=1.11×10^−85^), childhood intelligence (FDR=4.86 × 10^−27^), the number of fluid intelligence questions attempted (FDR=4.21×10^−13^), and fluid intelligence scores (FDR=2.23×10^−71^). In contrast, alcohol intake is negatively associated with cognitive performance (FDR=7.60 × 10^−45^), intelligence (FDR=2.78 × 10^−43^), childhood intelligence (FDR=1.24 × 10^−9^), fluid intelligence questions attempted (FDR=3.67 × 10^−10^), and fluid intelligence scores (FDR=4.58×10^−36^). These findings underscore the particularly strong influence of cheese and alcohol on cognitive health, with cheese showing the most robust positive correlation and alcohol the most significant negative correlation.

### Causal relationships between the consumption of dietary components and human intelligence

Given the correlations between multiple dietary components and cognitive functions, we further explored the causal relationships between these factors using two-sample MR (**Fig. 3A**). Our findings reveal that dried fruit consumption is positively associated with fluid intelligence (FDR=0.036), and both cooked vegetable (FDR=0.013) and coffee consumptions (FDR=0.037) with the number of fluid intelligence questions attempted. Remarkably, cheese consumption exhibited a positive causal association with all the traits, including cognitive performance (FDR=6.78×10^−12^), intelligence (FDR=6.43×10^−13^), childhood intelligence (FDR=9.09×10^−8^), and the number of fluid intelligence questions attempted (FDR=7.53×10^−6^) (**Fig.3A, Fig. S1**). Other MR methods including MR egger, weighted median, simple mode, and weighted mode all support the benefits of cheese consumption (**Fig. 3B-E, Fig. S1**). Conversely, alcohol consumption demonstrated a negative association with cognitive performance (FDR=9.67×10^−4^), intelligence (FDR=1.59×10^−3^), and the number of fluid intelligence questions attempted (FDR=0.01) (**Fig. 3A, Fig. S2**). Information of genetic instruments used in the analyses is detailed in **Table S2**. Sensitivity tests indicate that no individual instruments exerted undue influence on the overall causal estimate (**Table S3**). MR Egger regression intercept test revealed no evidence of pleiotropy (**Table. S4**).

**Fig. 3.**
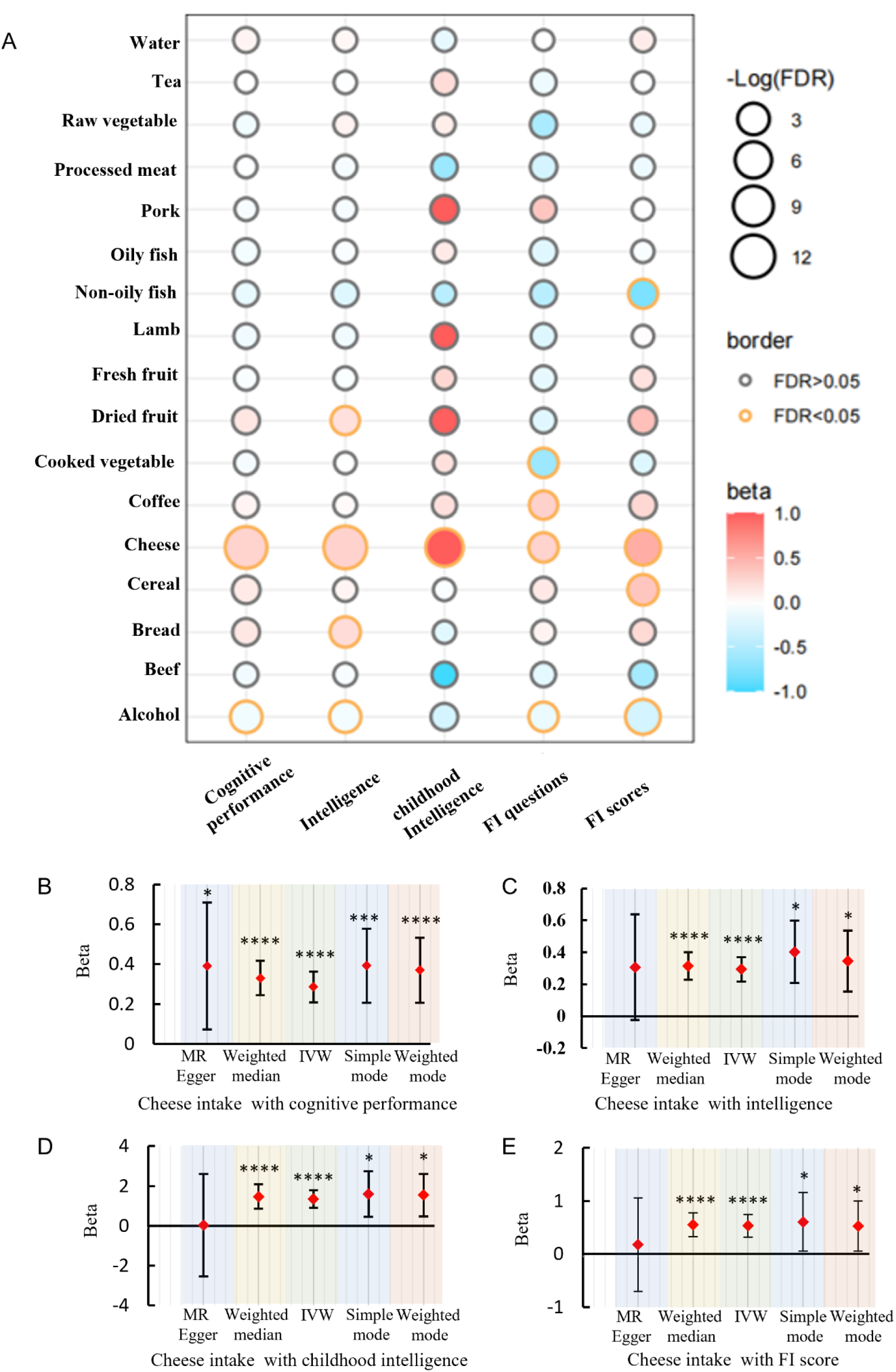
Causal relationships between dietary components and cognitive functions analyzed by two-sample MR. **A.** A bubble plot depicting the causal relationships between dietary components and cognitive functions. The color of the bubbles corresponds to effects size (beta) calculated by MR using the IVW method. The size of the bubbles corresponds to the negative logarithm of the association FDR. All tests were adjusted for multiple comparisons, and associations passing the threshold of FDR<0.05 are indicated by a yellow border. B-E forest plots depicting the causal estimates between cheese consumption and several cognitive functions by using five different MR methods.

Among all the dietary components studied, cheese and alcohol stand out, respectively, as having the strongest positive and negative associations with cognitive functions. To validate the causal effects of cheese, we used another large-scale genetic dataset for cheese consumption. Consistently, it still showed highly significant positive causal effects on all the traits examined (**Fig. S3**).

### Genome-wide meta-analysis identifies novel loci for cheese consumption

We performed a meta-analysis of GWAS for cheese consumption using two large-scale datasets and identified variants that reached genome-wide significance (p<5×10^−8^), as illustrated in **Fig. 4A**. The quantile-quantile (Q-Q) plot of the meta-analysis is presented in **Fig. S4A**. We analyzed association results for 5,660,731 genetic variants related to cheese intake. Among these, we observed 71 variants with genome-wide significant associations with cheese consumption, 34 of which were located more than 500 KB away from any previously reported variants (**Table S5**). We performed fine-mapping of these novel loci, as shown in **Fig. S4B**.

**Fig. 4.**
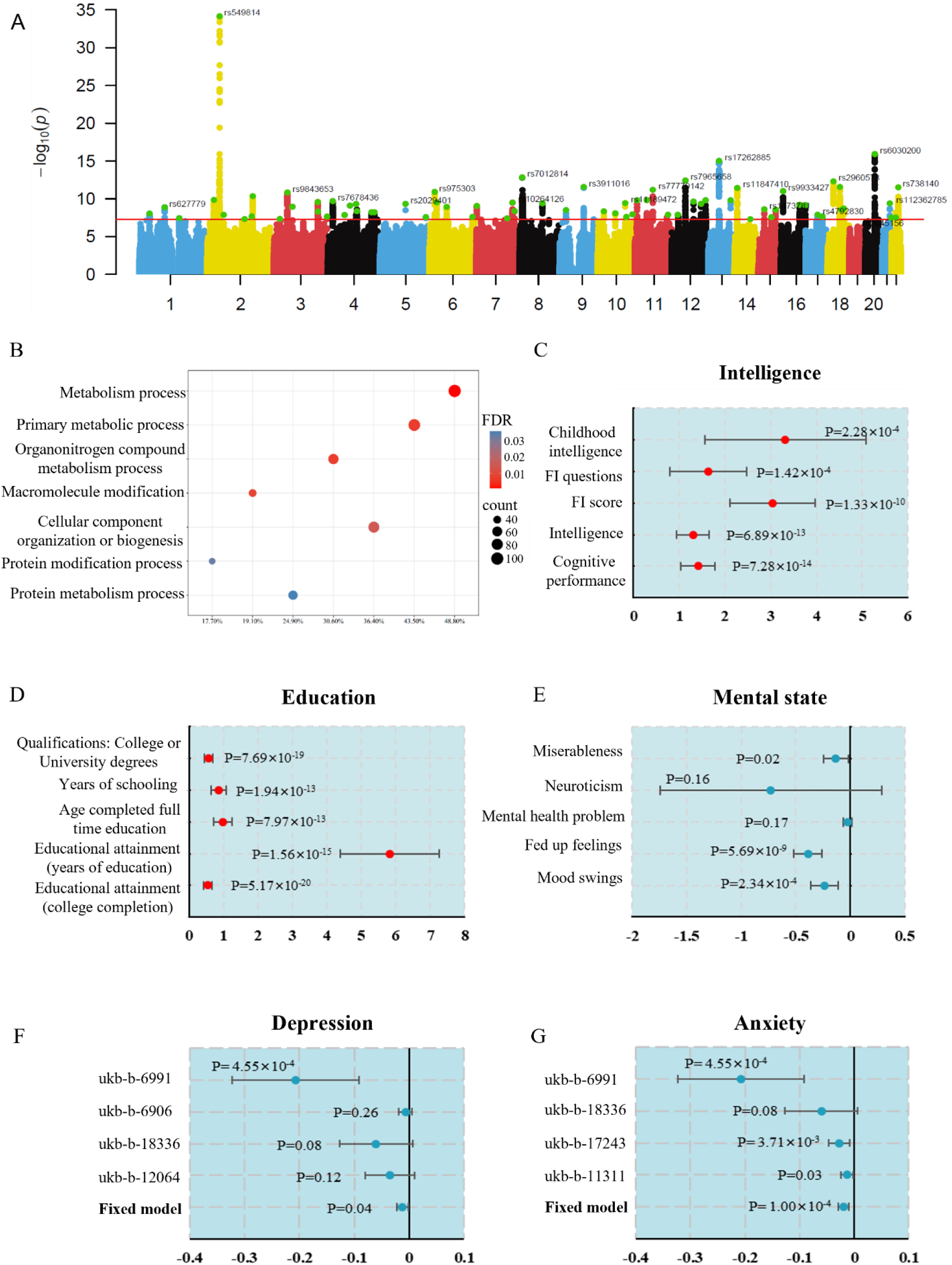
Meta-analysis of genome-wide association study (GWAS) for cheese consumption. **A** Manhattan plot showing the –log_10_(P value) of association for each SNP from the GWAS meta-analysis plotted on the y-axis against genomic position on the x-axis. The red dotted line corresponds to the genome-wide significance threshold (p<5×10^−8^). The clumped significant loci are highlighted and the top hit on each chromosome are labeled. **B** Pathway enrichment analysis using positional mapped genes from the meta-analysis recovers pathways relevant to cheese consumption. **C** Two-sample MR using newly identified genetic variables in the meta-analysis recovers the causal effects of cheese consumption on the cognitive functions. **D-G** Two-sample MR identifies causal effects of cheese consumption on extensive phenotypes including education (**D**), mental state (**E**), Depression (**F**), and anxiety (**G**). A fixed model was used for the meta-analysis of the MR results for depression and anxiety.

### Enrichment analysis reveal pathways relevant to cheese consumption

To gain insights into the biological processes responsible for cheese consumption, we performed gene pathway enrichment analysis. We analyzed the 71 significant variants using GTEx V8. These variants are associated with 1118 eQTLs (p<10^−4^), corresponding to a total of 247 unique genes (**Table S6-7**). After restricting the analysis to pathways described in Gene Ontology, KEGG, and Reactome, we observed 23 enriched pathways (FDR<0.05) (**Table S8**). Biological pathways associated with cheese consumption include organonitrogen compound metabolic process (FDR=7.20×10^−3^), macromolecule modification (FDR=0.04), protein modification process (FDR=0.03), and protein metabolic process (FDR=0.04) (**Fig. 4B**). These pathways possibly affect the metabolic process after cheese intake and influence the utilization of nutrients therein.

### Additional MR recovers the causal effects of cheese consumption on brain functions

Using the GWAS summary statistics after meta-analysis, we first confirmed the causal relationships between cheese intake and the five traits examined above by two-sample MR. (**Fig. 4C**). Next, we examined the potential effects of cheese on a broader range of related phenotypes. Interestingly, cheese consumption showed highly significant positive links with education related traits, including college or university degrees (p=7.69×10^−19^), years of schooling (p=7.69×10^−13^), and college completion (p=5.17×10^−20^) (**Fig. 4D**). Moreover, cheese also showed negative associations with phenotypes related psychological states, such as miserableness (p=0.02), mood swings (p=2.34×10^−4^), and feeling fed up (p=5.69×10^−9^) (**Fig. 4E**). Therefore, we further tested the causal effects of cheese consumption on depression and anxiety using multiple datasets and meta-analyzed the results. Consistently, the meta-analysis indicates that it shows a negative causal association with both depression (p=0.04) and anxiety (p=1.00×10^−4^) (**Fig. 4F-G**). Detailed information about the genetic instruments used in the analysis is shown in **Table S9**. MR Egger regression intercept test revealed no evidence of pleiotropy (**Table. S10**).

Additionally, we examined the causal effects of cheese consumption on 3143 traits related to brain region volumes described by Elliott et al. (*17*). No significant causal relationships were detected after adjustment for multiple comparisons (FDR<0.05) (**Table S11**). The volumes of brain regions are likely more determined by genetics rather than significantly influenced by dietary factors. Cheese may primarily enhance brain function rather than affecting its structures.

### eQTL-based MR indicates that cheese regulates cognition-involved genes in brain

To gain insights into the mechanisms through which cheese benefits intelligence, we explored the regulatory effects of cheese consumption on gene expression in twelve brain tissues using two-sample MR (**Fig. S5A**). The tissue-specific eQTLs were derived from GTEx. We could detect the regulatory effects of cheese in anterior cingulate cortex BA24, frontal cortex BA9, caudate basal ganglia, nucleus accumbens basal ganglia, cerebral cortex, putamen basal ganglia, spinal cord cervical, amygdala, cerebellum, cerebellar hemisphere, and hippocampus (FDR<0.05) (**Fig. S5B**). Some of the regulated genes, such as *LRRC37A* and *ARL17B*, have already been reported to be involved in brain structures and functions(*18*). Moreover, we also queried the knockout (KO) models for these genes in Mouse Genomics Informatics (MGI) and found genes relevant to brain functions (**Fig. S5C**). For example, KO of *GPR161*, regulated by cheese in caudate basal ganglia (FDR=0.02), results to abnormal morphology of the midbrain, spinal cord, and neural tube. KO of *ST8SIA3*, regulated in in frontal cortex BA9 (FDR=6.01×10^−4^), leads to abnormal locomotor behavior. KO of *SPTSSB*, regulated in hippocampus (FDR=5.77×10^−3^), causes optic nerve degeneration, brain vacuoles, abnormal cerebellum white matter morphology.

Additionally, we collected the genes regulated by cheese using a cut off of p<0.01 and obtained a list of 168 unique genes (**Table S12**). We performed protein-protein interaction (PPI) network analysis using the corresponding proteins encoded by these genes (**Fig. S5D**). This analysis revealed functional enrichment associated with only two human phenotypes (FDR<0.05): reaction time measurement (FDR=9.70×10^−3^) and cognitive function measurement (FDR=1.39×10^−2^). Collectively, these results suggest that cheese may regulate the expression of genes involved in cognitive functions in the brain.

### PheWAS analysis reveals the involvement of cheese-associated loci in brain functions

To explore the potential influence of the loci for cheese consumption on possible related phenotypes, we proceeded with a Phenome-wide association study (PheWAS) by accessing the Open IEU Project. The analysis using the 71 loci generated totally 3663 hits (p<5×10^−8^), approximately 18% of which are associated with brain and neurological functions related traits (**Table S13**). We observed concordant associations between some of the loci and traits related to brain morphology and functions. Of note, rs13107325, negatively associated with cheese consumption (p=4.88×10^−10^), had 351 hits (**Fig. S6A**). It is negatively associated with cognitive performance (p=1.09×10^−23^), intelligence (p=2.23×10^−21^), fluid intelligence score (ukb-a-5238, p=7.00×10^−16^), fluid intelligence score (ukb-a-5238, p=5.64×10^−15^), fluid intelligence questions attempted (p=3.60×10^−9^), educational attainment (p=8.80×10^−9^), college or university degrees (p=5.64×10^−8^), and years of schooling (p=4.45×10^−8^) (**Fig. S6B**). Additionally, we also observed its negative associations with neuroimaging intensity and contrast, measurements of brain regional volume and thickness, as well as other structural brain measurements (**Table S13**). The locus is located in the SLC39A8 region. SLC39A8 is a trace mineral transport gene, and it has been reported to affect brain development and nervous system function(*19*). The rs1014444, positively associated with cheese consumption (p=2.04 ×10^−8^), serves as another example. It is positively associated with cognitive performance (p=1.26×10^−9^), intelligence (p=5.45 ×10^−12^), years of schooling (p=1.80×10^−12^), and resting-state functional MRI connectivity (p=1.48×10^−11^) (**Fig. S6C**).

### Mouse knock-out models for positional mapped genes identified by GWAS

By positional mapping of the loci significantly associated with cheese consumption on human genome, we obtained a list of 76 genes (**Table S5**). To explore the roles that the genes play in the modulation of cognitive functions, we queried for KO mouse models using MGI. 44 out of the 76 genes were available in MGI (**Table S14**). The KO models indicate that more than a half of the available genes are associated with brain and neural functions. For example, KO models on *NPC1* (rs11665096) exhibited impaired learning, decreased brain size, and abnormal neuron morphology; KO of *ARID1B* (rs2078047) led to cognitive inflexibility, abnormal neuron morphology, and increased fear-related response; and KO of *SYT1* (rs6539284) resulted in abnormal synaptic transmission, abnormal neuron physiology, and abnormal excitatory postsynaptic currents (**Table 1**, **Table S14**).

**Table 1.**
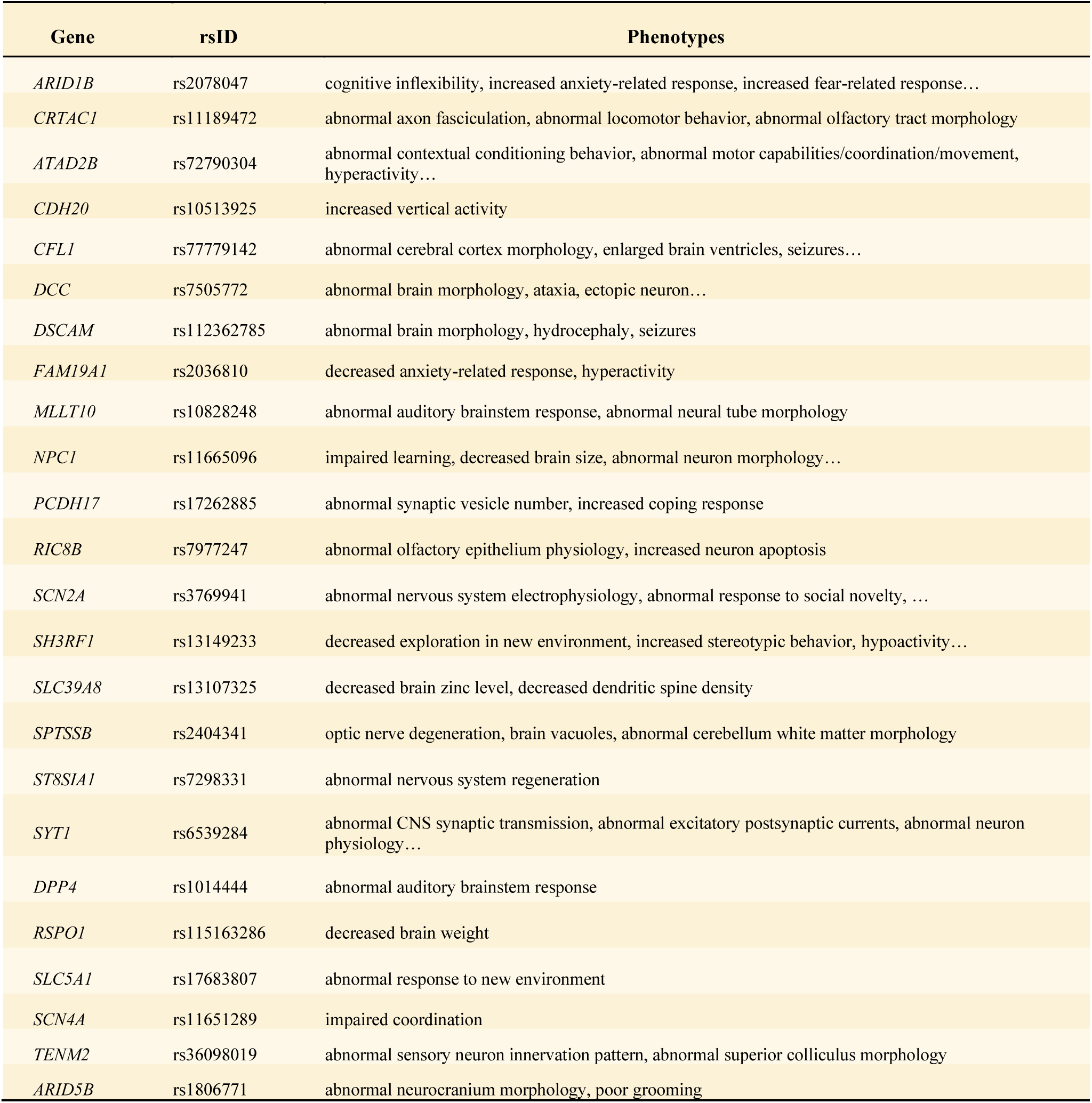
Knock-out of the positional mapped genes from the meta-analysis results in alterations to the structure and function of the brain and nervous system (information retrieved from Mouse Genome Informatics)

### PPI network analysis identifies cognition and intelligence as the leading enriched phenotypes

To explore human phenotypes linked to the loci related to cheese consumption, we performed PPI network analysis using genes derived from eQTL mapping. There are 186 nodes retained in the PPI network (**Fig. S7**). The proteins have more interactions among themselves (135 edges) than what would be expected for a random set of proteins of the same size and degree distribution drawn from human genome, with a PPI enrichment p=6.26 × 10^−11^. Functional enrichment analysis revealed 39 human phenotypes that were significantly enriched (FDR<0.05), with cognition (FDR=4.68×10^−19^) and intelligence (FDR=5.54×10^−17^) as the top two leading phenotypes (**Fig. S5A**, **Table 2**). Many other phenotypes that related to emotions, personality, and psychological and behavioral characteristics are also observed. These phenotypes include educational attainment (FDR=6.26 × 10^−8^), mental or behavioral disorder biomarker (FDR=5.78× 10^−6^), neuroticism measurement (FDR=6.38 × 10^−6^), personality trait measurement (FDR=1.36 × 10^−5^), feeling miserable measurement (FDR=4.53 × 10^−5^), mathematical ability (FDR=1.10 × 10^−4^), mood instability measurement (FDR=10^−3^), poor motor coordination (FDR=0.01), among others. Of note, we found approximately 40% of the significantly enriched phenotypes were related to brain functions (**Table 2**). Additionally, we also analyzed the genes derived from positional mapping of the loci associated with cheese consumption and observed similar results (**Fig. 5B**). These findings suggest that biological processes related to the metabolism of cheese-like foods potentially involve in cognitive functions.

**Fig. 5.**
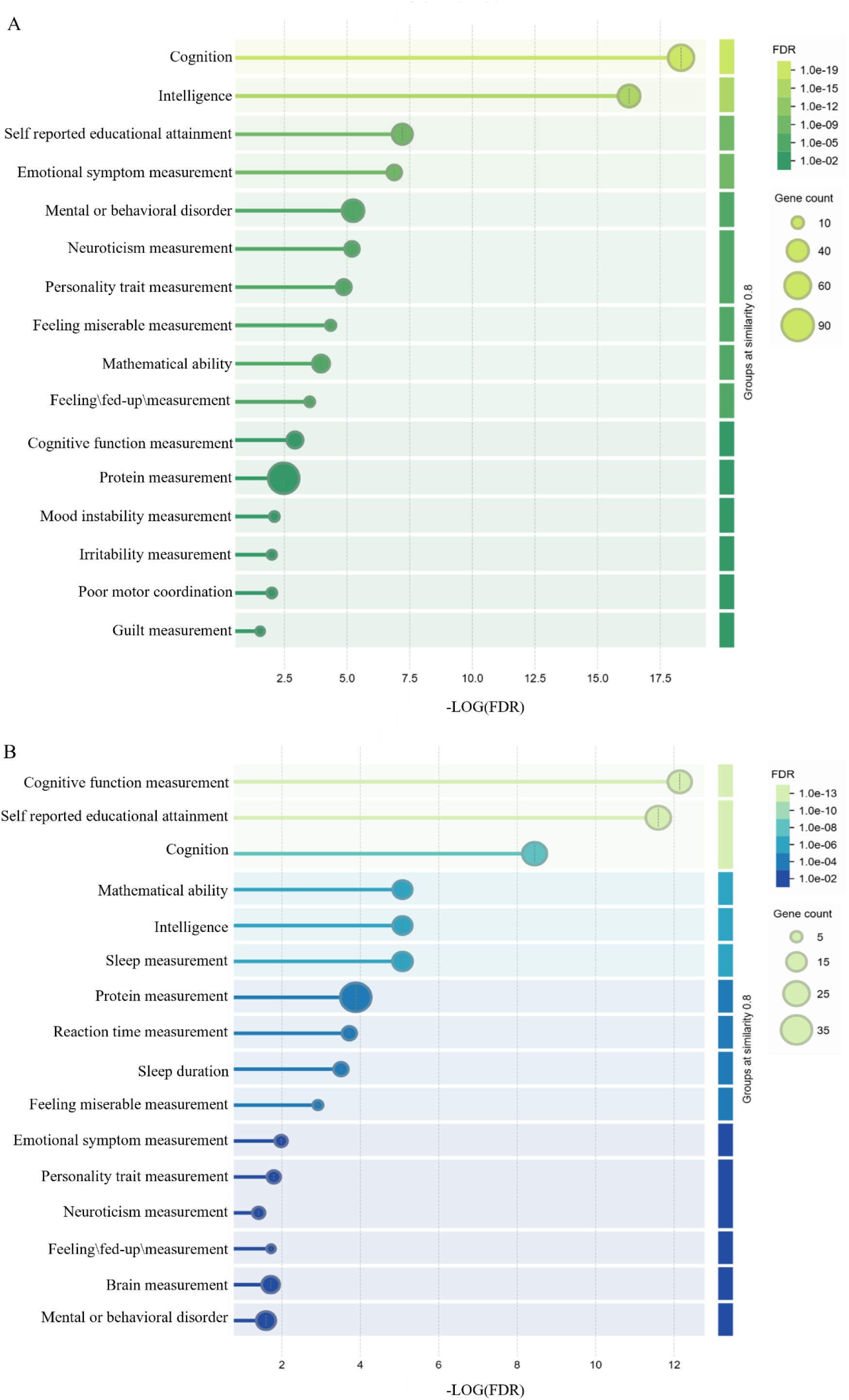
Functional enrichment analysis of protein-protein interaction network indicates that human phenotypes related to cognitive functions and intelligence are significantly enriched. The PPI network analysis was performed using the genes linked to the cheese consumption-associated loci identified by the meta-analysis of GWAS. These genes were identified by eQTL mapping (**A**) or positional mapping (**B**) of the loci to human genome. STRING was used to perform the enrichment analysis.

**Table 2.** Significantly enriched human phenotypes identified by protein-protein interaction network analysis of proteins encoded by e-QTL mapped genes of cheese consumption associated loci.

| Phenotype | Description | Count in network | Strength | FDR |
| --- | --- | --- | --- | --- |
| EFO:0003925 | **Cognition** | 58 of 1330 | 0.66 | 4.68E-19 |
| EFO:0004337 | **Intelligence** | 42 of 738 | 0.78 | 5.54E-17 |
| EFO:0004324 | Body weights and measures | 87 of 4122 | 0.35 | 1.54E-11 |
| EFO:0004302 | Anthropometric measurement | 88 of 4304 | 0.34 | 5.15E-11 |
| EFO:0004342 | Waist circumference | 37 of 1012 | 0.59 | 4.32E-09 |
| EFO:0004340 | Body mass index | 41 of 1257 | 0.54 | 6.61E-09 |
| EFO:0004784 | **Self-reported educational attainment** | 35 of 1012 | 0.56 | 6.26E-08 |
| EFO:0007803 | **Emotional symptom measurement** | 17 of 222 | 0.91 | 1.31E-07 |
| EFO:0009695 | Household income | 13 of 131 | 1.02 | 1.11E-06 |
| EFO:0007861 | Body ratio measurement | 32 of 1023 | 0.52 | 3.56E-06 |
| EFO:0006848 | **Mental or behaviuoral disorder biomarker** | 41 of 1615 | 0.43 | 5.78E-06 |
| EFO:0007660 | **Neuroticism measurement** | 17 of 298 | 0.78 | 6.38E-06 |
| EFO:0007911 | **Personality trait measurement** | 18 of 358 | 0.73 | 1.36E-05 |
| EFO:0007789 | BMI-adjusted waist circumference | 27 of 814 | 0.55 | 1.63E-05 |
| EFO:0004343 | Waist-hip ratio | 30 of 1006 | 0.50 | 2.36E-05 |
| EFO:0009598 | **Feeling miserable measurement** | 7 of 32 | 1.36 | 4.53E-05 |
| EFO:0004875 | **Mathematical ability** | 24 of 732 | 0.54 | 1.10E-04 |
| EFO:0008328 | Chronotype measurement | 16 of 334 | 0.71 | 1.20E-04 |
| EFO:0009588 | **Feeling \fed-up\ measurement** | 6 of 27 | 1.37 | 3.10E-04 |
| EFO:0007788 | BMI-adjusted waist-hip ratio | 24 of 780 | 0.51 | 3.10E-04 |
| EFO:0004732 | Lipoprotein measurement | 34 of 1426 | 0.4 | 3.50E-04 |
| EFO:0008354 | **Cognitive function measurement** | 21 of 673 | 0.52 | 0.0012 |
| EFO:0004747 | Protein measurement | 84 of 5856 | 0.18 | 0.0034 |
| EFO:0004612 | High density lipoprotein cholesterol measurement | 21 of 740 | 0.48 | 0.0040 |
| EFO:0005093 | Hip circumference | 24 of 923 | 0.44 | 0.0040 |
| EFO:0004512 | Bone measurement | 43 of 2295 | 0.30 | 0.0040 |
| EFO:0005105 | Lipid or lipoprotein measurement | 46 of 2526 | 0.29 | 0.0040 |
| EFO:0005671 | Smoking behaviour measurement | 19 of 649 | 0.49 | 0.0070 |
| EFO:0008475 | **Mood instability measurement** | 6 of 54 | 1.07 | 0.0080 |
| EFO:0009594 | **Irritability measurement** | 5 of 33 | 1.21 | 0.0101 |
| HP:0002275 | **Poor motor coordination** | 6 of 57 | 1.05 | 0.0101 |
| EFO:0004529 | Lipid measurement | 43 of 2400 | 0.28 | 0.0101 |
| EFO:0007877 | Risky sexual behaviour measurement | 6 of 59 | 1.03 | 0.0110 |
| EFO:0004870 | Sleep measurement | 22 of 868 | 0.43 | 0.0110 |
| EFO:0007862 | Reproductive behaviour measurement | 9 of 164 | 0.76 | 0.0124 |
| EFO:0004586 | Complete blood cell count | 47 of 2849 | 0.24 | 0.0262 |
| EFO:0009595 | **Guilt measurement** | 4 of 22 | 1.28 | 0.0295 |
| EFO:0008039 | BMI-adjusted hip circumference | 19 of 762 | 0.42 | 0.0423 |
| HP:0002079 | Hypoplasia of the corpus callosum | 14 of 462 | 0.51 | 0.0484 |

## Discussion

The relationship between diet and brain function is a complex and significant area of study within nutritional neuroscience. The brain requires a variety of nutrients to function optimally, and these nutrients largely come from our diet(*20*). Mounting evidence has revealed that cognitive function is not solely influenced by alterations in brain health but is also critically dependent on nutritional status(*21*). Given the foundational role nutrition plays in brain health, specific foods often receive attention for their potential cognitive benefits or detriments.

Dietary preferences are shaped by a multitude of factors such as biological, psychological, sociocultural, and environmental influences. Genetic factors play a significant role in determining taste preferences, such as preferences for sweet or bitter tastes, and food tolerances, such as lactose intolerance(*22*). Our analysis indicates that the consumption of commonly eaten foods is significantly affected by genetic factors (**Fig. 2B**). Certain genetic variants play a role in determining how frequently individuals consume a given dietary component, which, in turn, may impact brain function. Further analysis demonstrates that certain foods indeed exhibit significant genetic correlations with cognition-related phenotypes, including cognitive performance, intelligence, childhood intelligence, and fluid intelligence. Among the seventeen foods analyzed, cheese and alcohol displayed the strongest positive and negative correlations, respectively, with these phenotypes (**Fig. 2C**). Consistently, our two-sample MR analysis also suggests that cheese and alcohol have the strongest positive and negative causal relationships, respectively, with the phenotypes (**Fig. 3A**).

The previously reported impact of alcohol on cognitive decline supports our identification of a negative causal relationship between alcohol consumption and cognitive function(*23*). Given that cheese showed the most significant positive causal association, we conducted a meta-analysis of GWAS and identified 71 genetic variants linked to its consumption. We mapped these variants to eQTL-linked genes and performed pathway enrichment analysis. Biological processes including the organonitrogen compound metabolic process and protein metabolic process are significantly enriched. Given that cheese is a rich protein source, these processes likely explain the genetic variants’ impacts on cheese consumption.

By conducting MR with the newly identified variants, we could recover the causal effects of cheese intake on all the five previously examined phenotypes. Possessing higher cognitive abilities benefits the successful completion of educational pursuits. Consistently, we detected a positive causal relationship between cheese consumption and several traits related to education. Anxiety and depression are known to impact cognitive functions and intelligence(*24*). Interestingly, our analysis reveals a significant negative causal relationship between cheese intake and traits related to mood and emotion. To gain more insights into the mechanisms by which cheese consumption benefits cognitive function, we analyzed its regulatory effects on gene expression in twelve brain tissues. We recovered genes that have either been associated with cognitive functions in the literature or have been indicated by the MGI database to cause phenotypic alterations related to brain morphology and functions when knocked out.

On the other hand, we found that some of the genetic variants associated with cheese consumption are also involved in cognitive functions. There are three pieces of evidence supporting this conclusion. First, mouse KO models reveals that approximately a half of the loci mapped genes available in MGI play a role in brain morphology, neural functions, neurodevelopment, and behaviors. Second, PheWAS analysis indicates that some genetic variants are concordantly associated with both cheese consumption and phenotypes related to cognitive functions and brain characteristics. Third, PPI network analysis of proteins encoded by cheese eQTL mapped genes identifies cognition and intelligence as the two leading phenotypes enriched in the network. These results suggest that certain genetic variants affect both cheese consumption and cognitive function through as yet unidentified mechanisms. Some genetic variants may predispose individuals to prefer specific foods, such as cheese, and these variants could also influence cognitive processes. It is hypothesized that certain genetic variants may enhance the body’s ability to utilize the nutrients found in cheese, potentially improving cognitive function.

Although a few studies have already reported the connection between cheese intake and cognitive function, the evidence remains mixed. A randomized controlled trial noted a significant increase in brain-derived neurotrophic factor in older women with mild cognitive impairment after three months of cheese consumption(*25*), while another study analyzed two-year follow-up data and revealed no relationship between the cheese consumption with any changes in cognitive performance(*26*). The dietary habits and lifestyle factors of participants significantly influence cognitive functions, and these factors are difficult to effectively control in human studies.

Cheese is a dairy product made by coagulating the protein casein. The beneficial effects of peptides derived from casein on cognitive function have also been reported. For example, a tripeptide Met-Lys-Pro (MKP) derived from bovine casein significantly attenuated cognitive decline in a mouse model of Alzheimer disease(*27*). After orally administration, MKP can be absorbed into the plasma. A clinical trial reported that MKP intake could enhance spatial cognitive abilities in adults without dementia(*28*), which support our findings that cheese consumption potentially boost intelligence in both healthy adults and children (**Fig. 2C**). Cheese is also an important dietary source of tyrosine and tryptophan. Tyrosine is the precursor of dopamine and norepinephrine, and its supplementation has been suggested to reverse cognitive decline or enhance cognitive performance(*29*). Tryptophan is essential for the synthesis of serotonin and melatonin. Low levels of extracellular serotonin are associated with impaired memory consolidation, and both animal and human studies have shown that central serotonin can modulate a wide array of cognitive processes(*30, 31*). Furthermore, melatonin not only regulates circadian rhythms but also supports hippocampal neurogenesis and cognitive functions(*32*). In addition, cheese is rich in a variety of vitamins such as A, B, D, E, and K, and minerals including calcium, zinc, selenium, and phosphorus. Supplementation of these nutrients has also been reported to have beneficial effects on cognitive performance(*33*).

The strengths of the current analysis are multiple. First, we utilized a data-driven approach (LDSC and MR analyses) and identified cheese as the most effective dietary component on human intelligence from seventeen commonly consumed foods. Second, we assessed multiple phenotypes related to cognition and intelligence, and also examined broader phenotypes involved in education and psychological states. Cheese intake is biologically consistent with improvements across all these phenotypes, which enhances the biological credibility of our findings. Third, we conducted a meta-analysis on cheese intake using a large number of individuals. MR analysis with newly identified genetic variants recovered causal effects of cheese intake on all the phenotypes. Fourth, we identified genes significantly regulated by cheese in multiple brain tissues, some of which are already known to be involved in the neurological functions. Fifth, we used three approaches (MGI querying, PheWAS, PPI network) to confirm that some of the cheese-associated loci also impact cognitive functions.

The degree of credibility on the causality identified by MR depends on whether the MR assumptions are valid. We utilized a range of intelligence-related phenotypes alongside extensive additional phenotypes, including education, mental states, depression, and anxiety. This diversity in phenotypes makes confounding by LD unlikely. Additionally, we found that some genetic variants are significantly associated with both cheese intake and intelligence-related phenotypes. When conducting MR, we excluded instruments that strongly associate with the outcome to minimize the possibility of horizontal pleiotropy. Moreover, in our eQTL-based MR, there are only a few instruments available. The potential bias from each variable could affect the robustness of the model. Additionally, we used only European populations in this study. Future studies aim to include a broader range of ethnic and racial groups will provide chances to verify the applicability of the findings across different populations.

In conclusion, our comprehensive study integrating MR and genetic analyses underscores the intricate interplay between diet and genetic predispositions in influencing human intelligence. Especially, our findings support the positive impact of cheese consumption on cognition, highlighting the potential for targeted nutritional interventions to optimize cognitive well-being.

## Materials and Methods

### Data source and study design

The dietary components analyzed in this study were cereals, bread, raw vegetables, cooked vegetables, fresh fruits, dried fruits, processed meats, pork, beef, lamb, oily fish, non-oily fish, cheese, water, alcohol, coffee, and tea. The genetic data for these diet measurements were extract from the project of the IEU analysis of UK Biobank phenotypes. These datasets include a large-scale European population, ranging from 421,764 to 462,346 individuals, consisting of both males and females. Phenotypes related to cognition included were cognitive performance, intelligence, childhood intelligence, fluid intelligence, and fluid intelligence questions attempted within time limit. Extensive phenotypes including educational attainment (college completion and years of education), mood swings, and Miserableness. Detailed information on these datasets is provided in **Table S1**.

### LD score regression

LD score regression analysis was performed using GWAS summary statistics to explore the inheritability of the consumption of various dietary components and the genetic correlation between these dietary intakes and phenotypes related to human intelligence. Briefly, the linkage disequilibrium (LD) score was calculated for each SNP. The 1000 Genomes Project was used as the LD reference panel for estimating the LD score. Each SNP’s statistic from GWAS was then regressed against its LD score using a regression model. Heritability of the dietary intakes was estimated from the regression coefficients. Additionally, the genetic correlation between two traits of interested were estimated by performing a regression analysis on the GWAS results of both traits, using the LD score as the explanatory variable. These computations were conducted using R (version 4.4.1) and the GenomicSEM package (version 0.0.5).

### MR on dietary consumptions and cognitive functions

#### Instrument selection

Genetic variants as instruments are selected based on the following criteria: (1) significantly associated with exposure, with a p-value < 5×10^−8^; (2) not strongly associated with the outcome, with a p-value > 10^−4^; (3) an LD threshold of R^2^<0.01 within a window of 5000 kb; (4) with an F-value > 10.

#### MR analysis

The TwoSampleMR package (version 0.5.7) was utilized. Inverse Variance Weighted (IVW), MR Egger, Weighted Median, Simple Mode, and Weighted Mode are used to estimate the causalities. The IVW method served as the primary analytical approach due to its efficiency when all instruments are valid. In case of detected heterogeneity among the instruments, a mixed random effects model was applied to account for the variability in the causal estimates. Otherwise, a fixed effect model was preferred to provide a precise estimate of the causal effects.

#### Sensitivity analysis

Heterogeneity among the instruments was evaluated using Cochran’s Q statistic for both the MR Egger and IVW methods. The MR Egger regression intercept test was employed to detect potential pleiotropy. We also conducted a leave-one-out sensitivity analysis to ensure robustness of the results. This approach iteratively excludes each variant from the analysis to examine the influence of individual variants on the overall causal inference.

### Genome-wide association analysis for cheese consumption

We performed a fixed-effect model meta-analysis on two large GWAS datasets from the IEU open GWAS project (ukb-b-1489) and the GWAS Catalog (GCST90096906). We first checked that summary statistics from GWAS are in a homogeneous format using R packages MungeSumstats (version 1.12.0). The meta-analysis was performed using R package metafor (version 4.6.0). Only SNPs present in both datasets were included in the analysis. The meta-analysis included a total of 817,328 individuals of European ancestry, with the objective of investigating the genetic factors influencing cheese consumption. We used FUMA to annotate the results using the default settings. SNPs that showed significant associations with cheese consumption (p < 5×10^−8^) were clumped within a 10,000 kb window using an R^2^ threshold of 0.001. The left SNPs that located more than 500 kb away from previously reported loci were identified as novel loci. The closest genes corresponding to each locus were retrieved using human genome HG19/GRCh37 as a reference.

### Pathway enrichment analysis

We performed an enrichment analysis to explore biological pathways linked to the loci associated with cheese consumption that met GWAS significance thresholds. For each locus, we pinpointed the leading variant (within a 1Mb region) and then identified eQTLs from GTEx V8 in all tissues associated with the top variants and extracted all genes with a p < 10^−4^. We compiled these genes into a unified gene set and performed pathway enrichment analysis using the Database for Annotation, Visualization and Integrated Discovery (DAVID) tool against the Gene Ontology, KEGG, and Reactome database. Pathways significantly enriched at an FDR < 0.05 were extracted and displayed.

### Two-sample MR on cheese consumption and brain region volumes

We performed two-sample MR on the consumption of cheese and 3143 traits related to brain region volumes. The specific collection of GWAS datasets (ubm-a) for the imaging phenotypes related to brain region volumes provided by UK Biobank were downloaded from the Open IEU GWAS project (https://gwas.mrcieu.ac.uk)(17). All the MR procedures are the same as those described above. The results of the IVW method were adjust for multiple comparisons using the Benjamin-Hochberg (BH) procedure.

### eQTL-based MR on cheese consumption and gene expression in multiple brain tissues

We inferred the causal relationships between cheese intake and gene expressions in twelve brain tissues including cerebral cortex, frontal cortex BA9, anterior cingulate cortex BA24, amygdala, caudate basal ganglia, putamen basal ganglia, nucleus accumbens basal ganglia, hippocampus, cerebellar hemisphere, cerebellum, substantia nigra, spinal cord cervical c-1 by two-sample MR analysis. The instruments were selected from the significant SNPs derived from the meta-analysis, and also significant SNPs that only exist in one dataset. The criteria for instrument selection were similar to those described above, except that the association significance was loosened (p < 10^−5^) to ensure we have enough instruments. We obtained tissue specific eQTLs for gene expressions from GTEx. IVW was used as the MR analytical method. The results were adjusted for multiple tests using the BH procedure.

### Querying the MGI database

We queried the Mouse Genome Informatics (MGI, https://www.informatics.jax.org/) resource for the positional mapped genes in the meta-analysis of GWAS for cheese consumption. The phenotypic alterations resulting from gene knockouts were retrieved. We manually curated the details for models that displayed abnormalities in the brain and nervous system.

### PheWAS analysis of the genetic variants associated with cheese consumption

Using the 71 significant variants identified in the meta-analysis of GWAS for cheese consumption, we conducted a PheWAS analysis by accessing the IEU Open GWAS Project via application programming interface (API). Phenotypes associated with any of the variants showing a p <5×10^−8^ were considered significant and extracted.

### Protein-protein interaction network analysis

The proteins encoded by cheese regulated genes in the brain and the cheese loci mapped genes from the meta-analysis, respectively, were utilized to conduct a protein-protein interaction (PPI) network analysis via the STRING database (https://string-db.org/). Both functional and physical protein associations were taken into consider. Whole genome was used as the statistical background for the functional enrichments in the network. Human phenotypes significantly enriched with a significance of FDR<0.05 were extracted.

### Statistical Analysis

Significance levels for associations between genetic variants and traits were set at p < 5×10^−8^. In MR analysis, significance was determined at IVW p < 0.05, with pleiotropy considered non-significant at p > 0.05. When multiple testing was applied, the false discovery rate (FDR) was calculated using the Benjamini-Hochberg (BH) method, with a significance threshold set at FDR < 0.05. The same FDR threshold was applied to determine significance in enrichment analysis.

## Data Availability

All data produced in the present study are available upon reasonable request to the authors

## Acknowledgments

We acknowledge all participants and staff involved in the UK Biobank and other related projects for their valuable contributions to the GWAS datasets utilized in this research.

## Data and materials availability

All data needed to evaluate the conclusions in the paper are present in the paper and/or the Supplementary Materials. All the GWAS summary data used in this study can be downloaded from the IEU open GWAS project (https://gwas.mrcieu.ac.uk/) and GWAS Catalog (https://www.ebi.ac.uk/gwas/).

## Supplementary Materials

Figure S1-S7

Table S1-S14

