## Supplementary figures for "Integration of Mendelian Randomization and Genome-wide Association Analysis Reveals Potential Benefits of Cheese Intake on Human Intelligence"

Peng Chen *et al.*

**This PDF file includes:**

Figs. S1 to S7

Tables S1 to S14

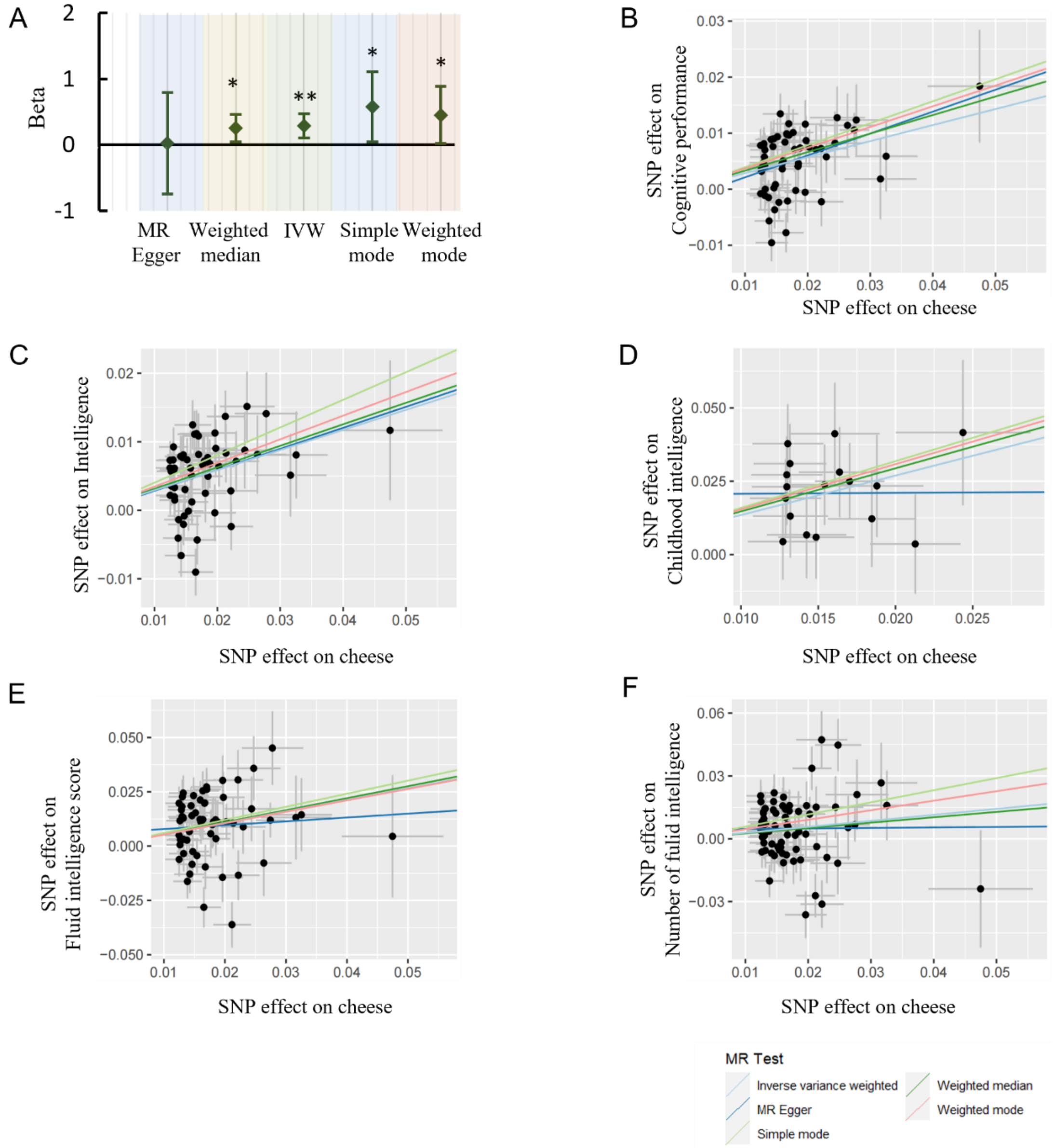


**Fig. S1 The causal relationships between cheese consumption and cognitive functions estimated by two-sample MR analysis**. **A**. A forest plot showing the causal effects (beta with 95% confidence interval) of cheese intake on fluid intelligence question attempted within time limit (*p<0.05, **p<0.01). **B-F**. Scatterplots showing the causal relationships between cheese intake and cognitive performance (**B**), intelligence(**C**), childhood intelligence (**D**), fluid intelligence score(**E**), and number of fluid intelligence questions attempted within time limit (**F**).

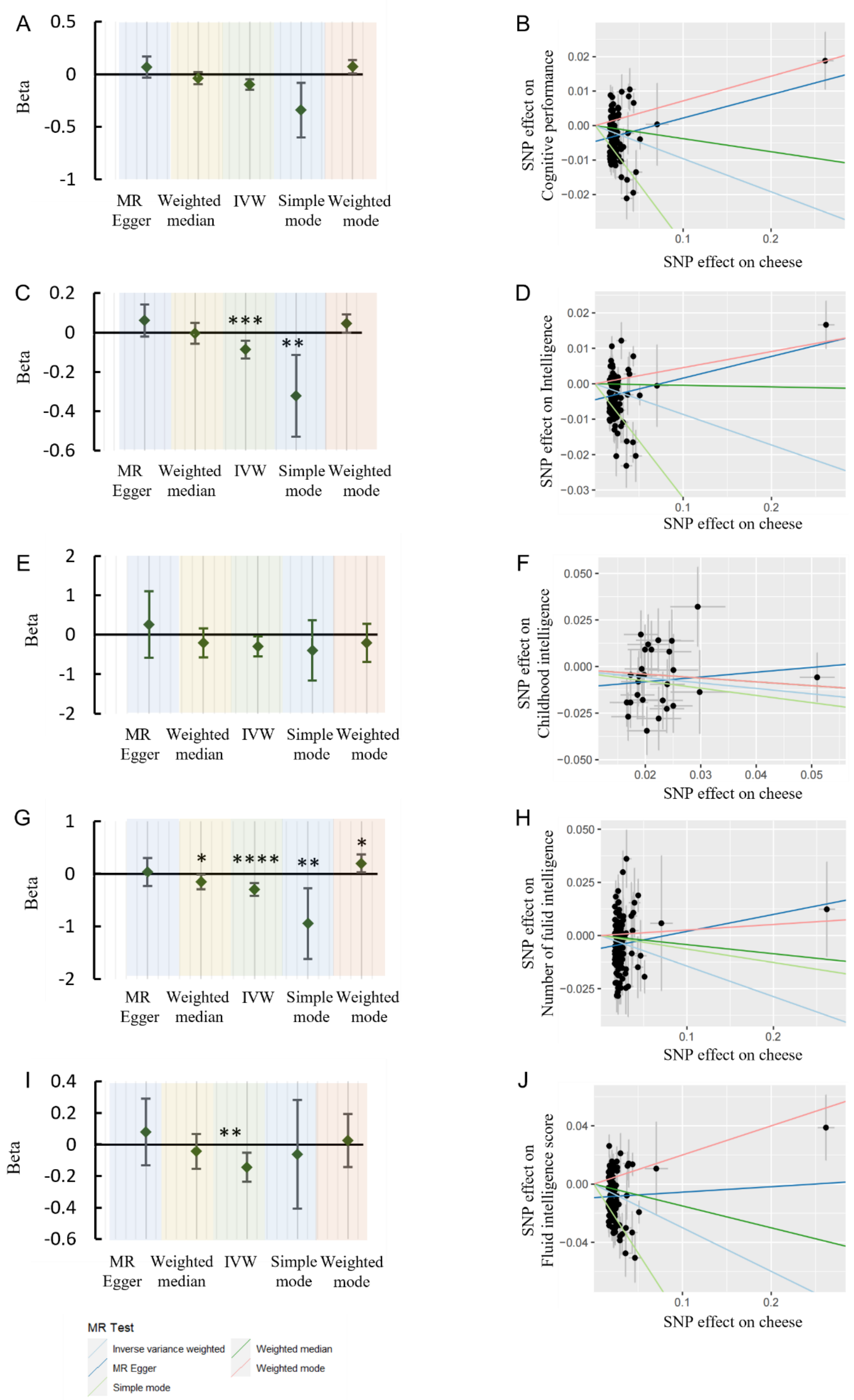
A
B
C
D
E
F
G
H
I
J
Beta
MR Egger
Weighted median
IVW
Simple mode
Weighted mode
SNP effect on Cognitive performance
SNP effect on Intelligence
SNP effect on Childhood intelligence
SNP effect on Number of fulid intelligence
SNP effect on Fluid intelligence score
SNP effect on cheese
MR Test
Inverse variance weighted
MR Egger
Simple mode
Weighted median
Weighted mode

**Fig. S2 The causal effects of alcohol consumption on cognitive functions evaluated by two-sample MR.** Forest plots (left panel) and scatterplots (right panel) showing the causal estimates of alcohol on cognitive performance (**A-B**), intelligence (**C-D**), childhood intelligence (**E-F**), numbers of fluid intelligence scores attempted within time limit (**G-H**), and fluid intelligence scores (**I-J**). (*p<0.05, **p<0.01, ***p<0.001, ****p<0.0001).

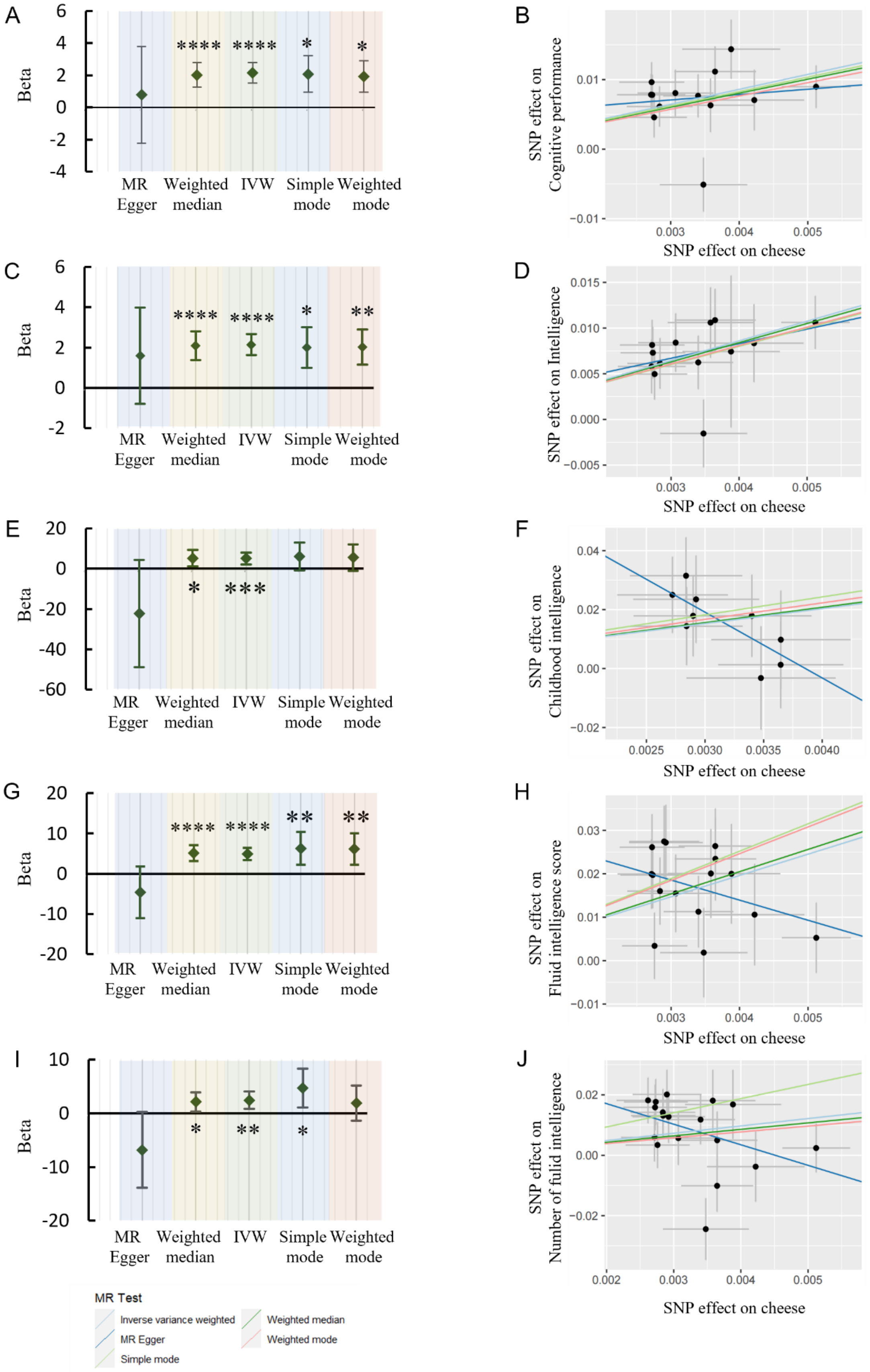
A
Beta
MR Egger
Weighted median
IVW
Simple mode
Weighted mode
B
SNP effect on Cognitive performance
SNP effect on cheese
C
D
SNP effect on Intelligence
E
F
SNP effect on Childhood intelligence
G
H
SNP effect on Fluid intelligence score
I
J
SNP effect on Number of fulid intelligence
MR Test
Inverse variance weighted
MR Egger
Simple mode
Weighted median
Weighted mode

**Fig. S3 Two-sample MR using another large-scale dataset for cheese consumption confirms the causal effects of cheese on cognitive functions**. Validating the detrimental effects of cheese consumption and brain-function related phenotypes. Forest plot depicting the causal effects (beta with 95% confidence interval) and Scatterplots depicting the causal relationships between cheese consumption and cognitive performance (**A**, **B**), number of intelligence (**C**, **D**), and childhood intelligence (**E**, **F**), and FI score (**G**, **H**), number of fluid intelligence questions attempted within time limit (**I**, **J**), (*p<0.05, **p<0.01, ***p<0.001, ****p<0.0001).

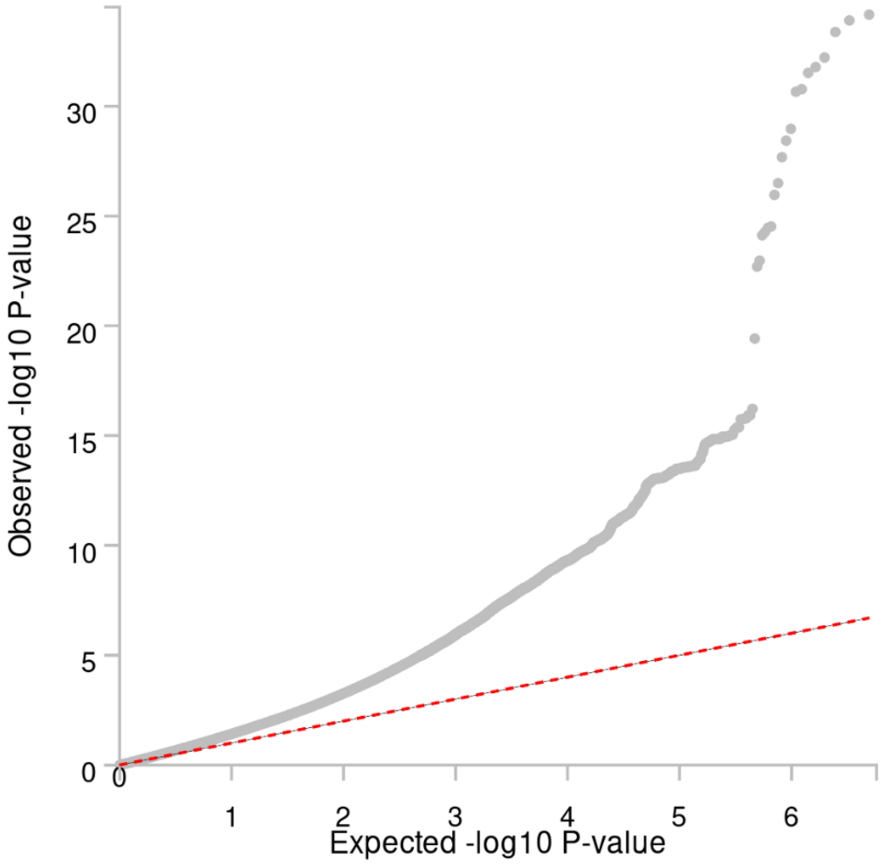


**Fig. S4A The quantile-quantile (Q-Q) plot of the meta-analysis for cheese consumption**.

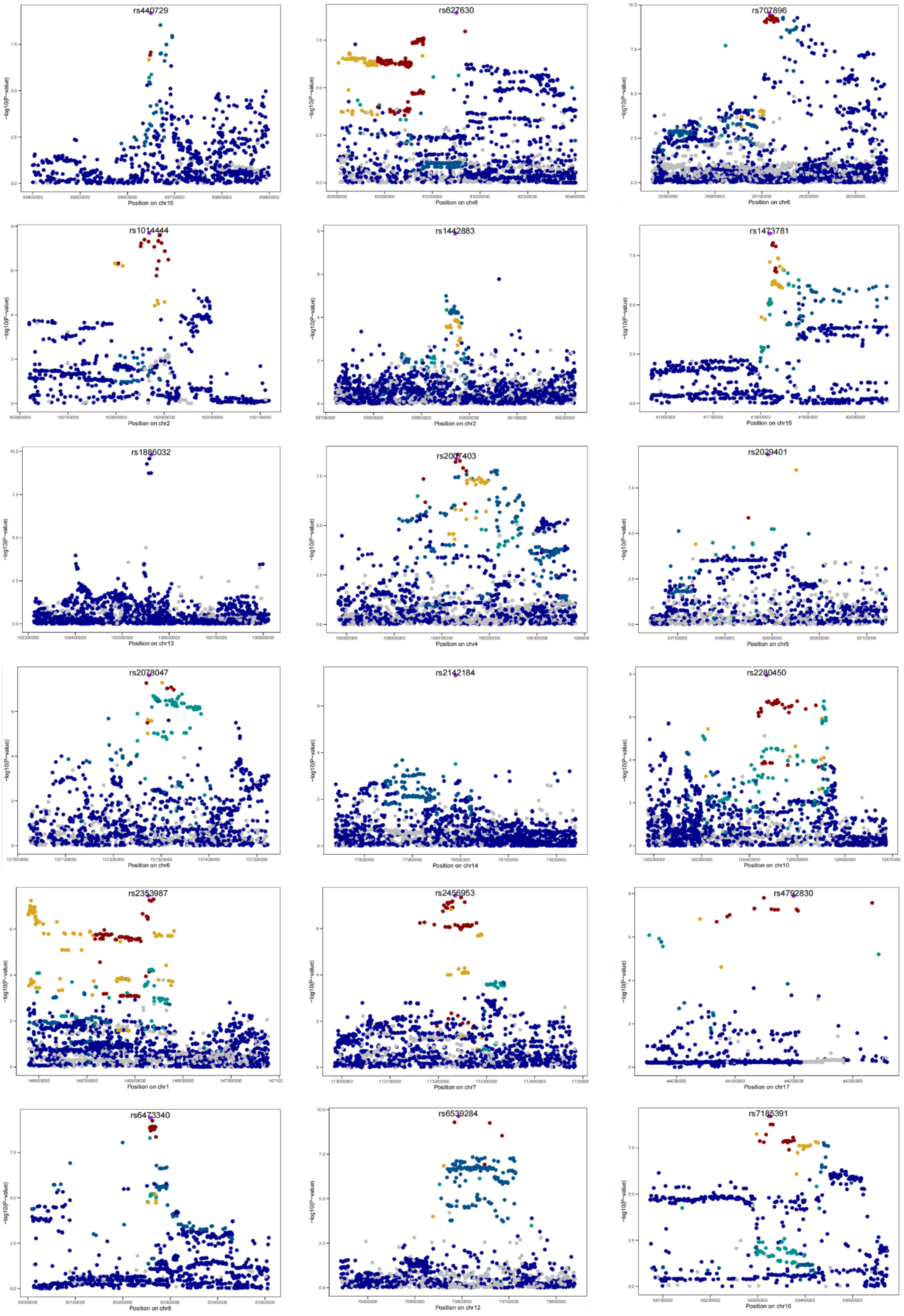
rs440729
Position on chr16
rs627630
Position on chr6
rs707896
Position on chr6
rs1014444
Position on chr2
rs1442883
Position on chr2
rs1473781
Position on chr15
rs1886032
Position on chr13
rs2007403
Position on chr4
rs2029401
Position on chr5
rs2078047
Position on chr6
rs2142184
Position on chr14
rs2280450
Position on chr10
rs2353987
Position on chr1
rs2456953
Position on chr7
rs4792830
Position on chr17
rs6473340
Position on chr8
rs6539284
Position on chr12
rs7185391
Position on chr16
-log10(P-value)

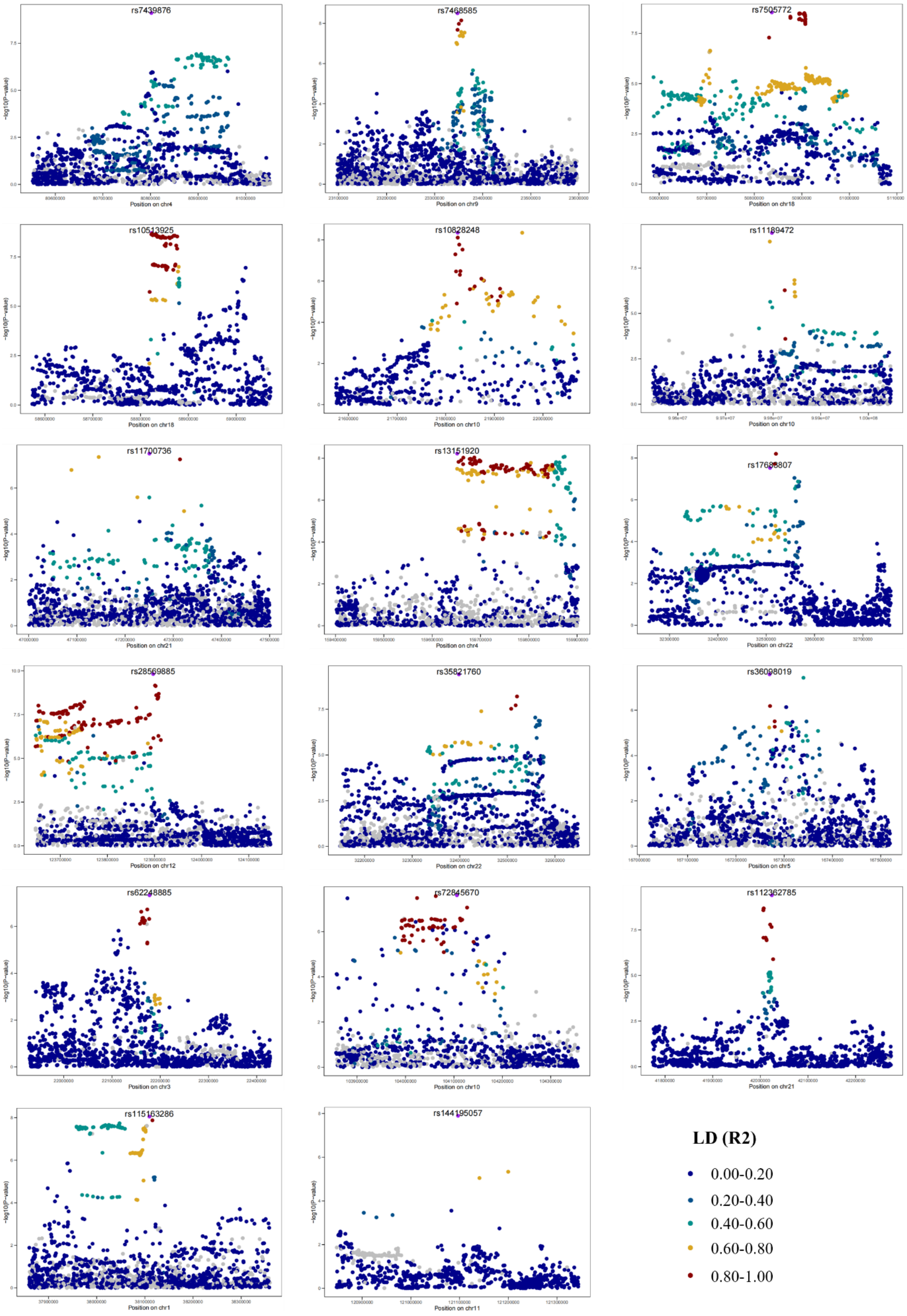

rs7439876
Position on chr4
rs7468585
Position on chr9
rs7505772
Position on chr18
rs10513925
Position on chr18
rs10828248
Position on chr10
rs11189472
Position on chr10
rs11700736
Position on chr21
rs13151920
Position on chr4
rs17683807
Position on chr22
rs28569885
Position on chr12
rs35821760
Position on chr22
rs36098019
Position on chr5
rs62248885
Position on chr3
rs72845670
Position on chr10
rs112362785
Position on chr21
rs115163286
Position on chr1
rs144195057
Position on chr11
-log10(P-value)
LD (R2)
0.00-0.20
0.20-0.40
0.40-0.60
0.60-0.80
0.80-1.00

**Fig. S4B Fine-mapping of the novel loci for cheese consumption identified in the meta-analysis of GWAS.**

A

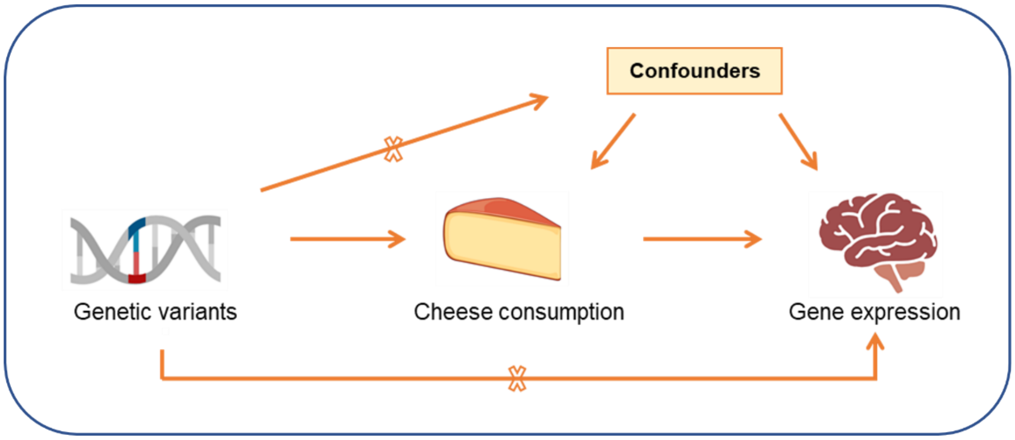


B

| Tissue | # | Gene |
|---|---|---|
| Anterior_cingulate_cortex_BA24 | 6 | *ARHGAP27*; *ARL17B*; *KANSL1*; *LRRC37A*; *NSFP1*; *RP1-240B8.3* |
| Frontal_cortex_BA9 | 6 | *C18orf8*; *KANSL1*; *LRRC37A*; *NSFP1*; *RP1-240B8.3*; *ST8SIA3* |
| Caudate_basal_ganglia | 5 | *ARL17B*; *GPR161*; *KANSL1*; *LRRC37A*; *NSFP1* |
| Nucleus_accumbens_basal_ganglia | 5 | *ARL17B*; *ID4*; *IL17RE*; *LRRC37A*; *NSFP1* |
| Cortex | 4 | *ARL17B*; *LRRC37A*; *NSFP1*; *RP1-240B8.3* |
| Putamen_basal_ganglia | 4 | *ARL17B*; *KANSL1*; *LRRC37A*; *NSFP1* |
| Spinal_cord_cervical_c-1 | 4 | *AC009299.3*; *LRRC37A*; *NSFP1*; *PCDP1* |
| Amygdala | 3 | *ARL17B*; *LRRC37A*; *NSFP1* |
| Cerebellum | 2 | *ARL17B*; *NSFP1* |
| Cerebellar_Hemisphere | 1 | *NSFP1* |
| Hippocampus | 1 | *SPTSSB* |
| Substantia_nigra | 0 | |

C

| Genes | Name | Feature Type | Term |
|---|---|---|---|
| *GPR161* | G protein-coupled receptor 161 | protein coding gene | abnormal midbrain morphology<br>abnormal spinal cord morphology<br>abnormal neural tube morphology<br>abnormal midbrain development<br>abnormal neural fold formation<br>increased midbrain size<br>increased neuronal precursor proliferation |
| *ST8SIA3* | ST8 alpha-N-acetyl-neuraminide alpha-2,8-sialyltransferase 3 | protein coding gene | abnormal locomotor behavior |
| *SPTSSB* | serine palmitoyltransferase, small subunit B | protein coding gene | optic nerve degeneration<br>brain vacuoles<br>abnormal cerebellum white matter morphology |
| *ID4* | inhibitor of DNA binding 4 | protein coding gene | decreased brain size<br>abnormal stratification in cerebral cortex<br>abnormal hippocampus development<br>abnormal brain development<br>abnormal forebrain development<br>premature neuronal precursor differentiation<br>abnormal basal ganglion morphology<br>abnormal neocortex morphology<br>abnormal neuron differentiation |

D

| Phenotype | Description | Count in network | Strength | FDR |
|---|---|---|---|---|
| EF0:0008393 | Reaction time measurement | 10 of 235 | 0.85 | 0.0139 |
| EF0:0008354 | Cognitive function measurement | 17 of 673 | 0.62 | 0.0097 |

**Fig. S5 Two-sample MR identifies genes significantly regulated by cheese in multiple brain tissues**. **A** Diagram depicting the analysis model. **B** genes significantly regulated by cheese in each tissue (FDR<0.05)**. C** Knockout mouse models indicate that some of the cheese regulated genes are involved in brain morphology and functions. **D** Protein-protein interaction network analysis of the proteins encoded by the genes that regulated by cheese (p<0.01) reveals the significant enrichment of cognitive function related phenotypes.

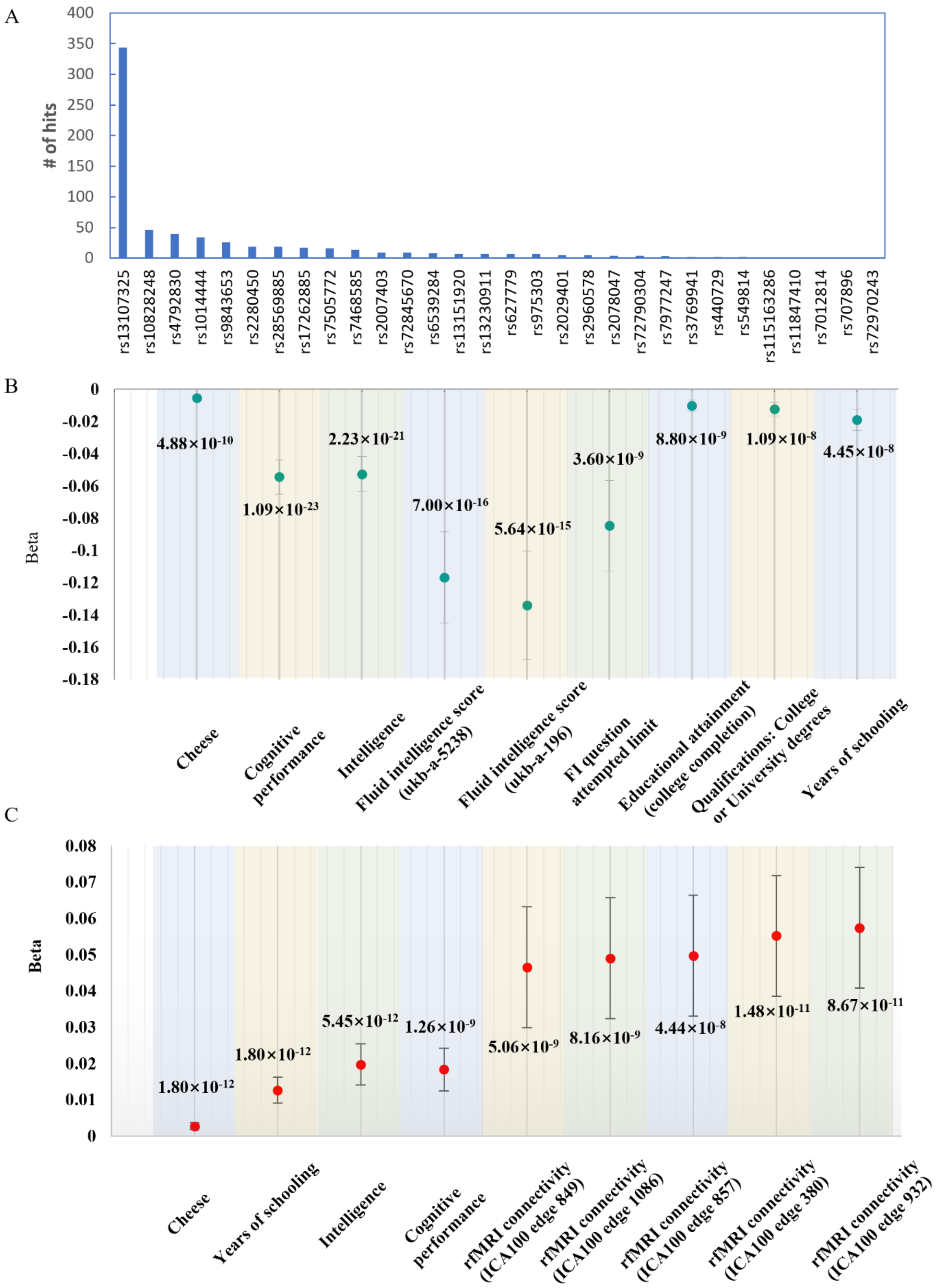
A
### of hits
400
350
300
250
200
150
100
50
0
rs13107325
rs10828248
rs4792830
rs1014444
rs9843653
rs2280450
rs28569885
rs17262885
rs7505772
rs7468585
rs2007403
rs72845670
rs6539284
rs13151920
rs13230911
rs627779
rs975303
rs2029401
rs2960578
rs2078047
rs72790304
rs7977247
rs3769941
rs440729
rs549814
rs115163286
rs11847410
rs7012814
rs707896
rs72970243
B
Beta
0
-0.02
-0.04
-0.06
-0.08
-0.1
-0.12
-0.14
-0.16
-0.18
4.88×10-10
1.09×10-23
2.23×10-21
7.00×10-16
5.64×10-15
3.60×10-9
8.80×10-9
1.09×10-8
4.45×10-8
Cheese
Cognitive performance
Intelligence
Fluid intelligence score (ukb-a-5238)
Fluid intelligence score (ukb-a-196)
FI question attempted limit
Educational attainment (college completion)
Qualifications: College or University degrees
Years of schooling
C
Beta
0.08
0.07
0.06
0.05
0.04
0.03
0.02
0.01
0
1.80×10-12
1.80×10-12
5.45×10-12
1.26×10-9
5.06×10-9
8.16×10-9
4.44×10-8
1.48×10-11
8.67×10-11
Cheese
Years of schooling
Intelligence
Cognitive performance
rfMRI connectivity (ICA100 edge 849)
rfMRI connectivity (ICA100 edge 1086)
rfMRI connectivity (ICA100 edge 857)
rfMRI connectivity (ICA100 edge 380)
rfMRI connectivity (ICA100 edge 932)

**Fig. S6 Phenome-wide association analysis of the significant loci for cheese consumption**. **A** Number of hits of each locus. **B-C** Association of rs13107325 (**B**) and rs1014444 (**C**) with phenotypes related to brain functions.

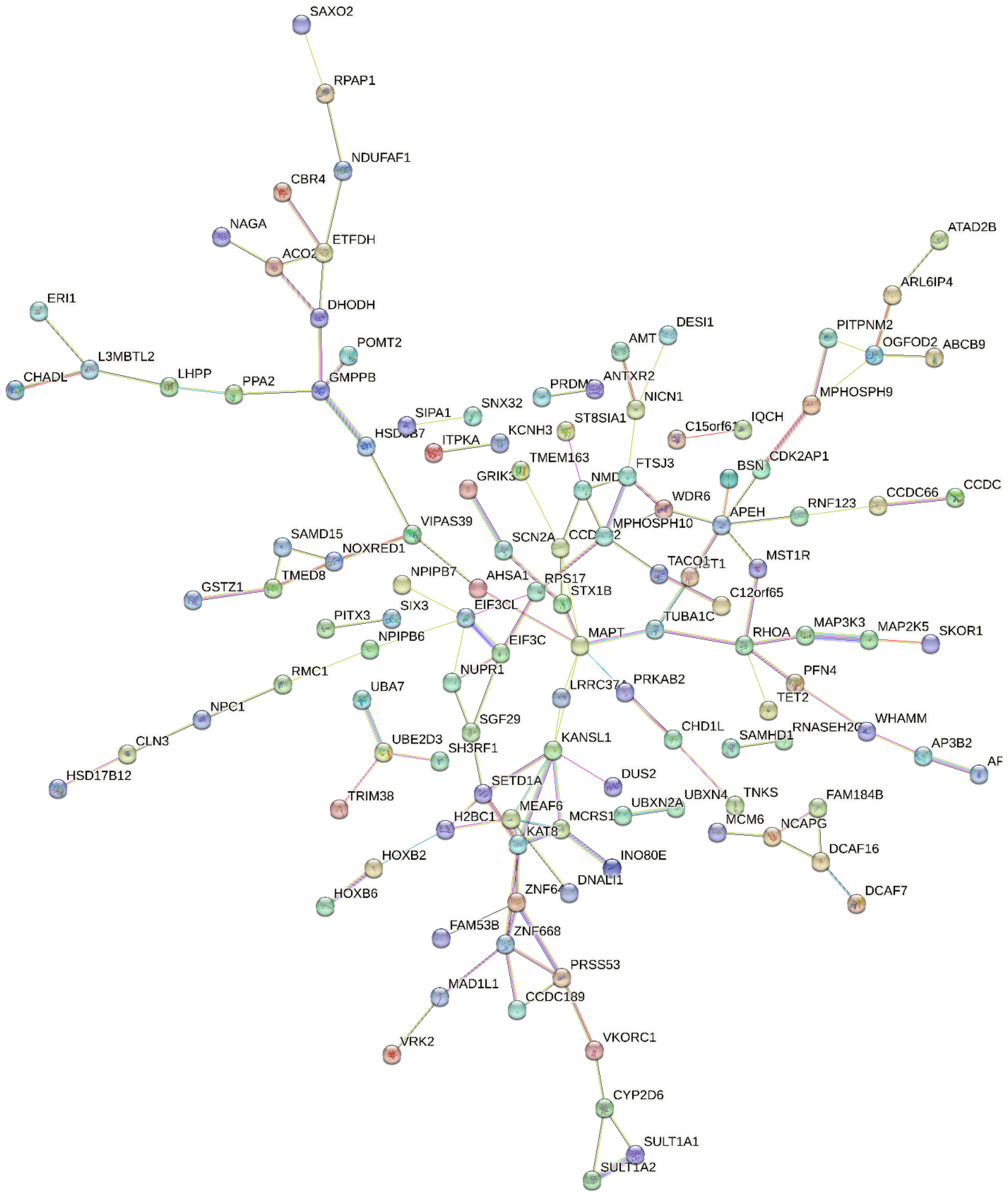


**Fig. S7 Protein-protein interaction network analysis of the proteins encoded by cheese eQTLs mapped genes**. Full network was exported from STRING (version 12.0). The edges indicate both functional and physical protein associations. The minimum required interaction score was set at 0.4. Disconnected nodes are hidden in the network.

**Legends for Table S1-S13 (Separate file)**

Table S1: GWAS datasets and related information used in the present research.

Table S2: Instrumental variables and their associated statistics used in the MR analyses on the consumption of dietary components and cognition-related phenotypes.

Table S3: sensitivity test using the "leave-one-out" approach for the MR analyses on the consumption of dietary components and cognition-related phenotypes.

Table S4: Results of Mendelian randomization analysis on the causal relationships between different dietary components and intelligence-related phenotypes, along with tests for heterogeneity and pleiotropy.

Table S5: Genes identified by positional mapping of the loci significantly associated with cheese consumption onto human genome.

Table S6: Number of cis-eQTLs per GTEx V8 tissue from previously published and newly identified cheese intake genome-wide significant loci.

Table S7: The eQTL mapping of the significant loci identified by meta-analysis of GWAS for cheese consumption ($p<1E-4$)

Table S8: Significant pathways identified by enrichment analysis using cheese loci eQTL linked genes ($FDR<0.05$).

Table S9: Statistics of the instrumental variants for assessing causal relationships between cheese intake and brain function-related phenotypes.

Table S10: Results of Mendelian randomization analysis on the causal relationships between cheese intake and brain function-related phenotypes, along with tests for heterogeneity and pleiotropy.

Table S11: The results of two-sample Mendelian randomization analysis on cheese consumption and 3143 traits related to brain region volumes.

Table S12: eQTL-based MR identify genes regulated by cheese consumption in twelve brain tissues ($p<0.01$).

Table S13: Phenome-wide association study (Phe-WAS) of the significant loci for cheese consumption.

Table S14: Phenotypes of knockout models of genes identified by meta-analysis of GWAS for cheese consumption, retrieved in the Mouse Genome Informatics (MGI) database.
